# Functional Connectivity and Complexity Analyses of Resting-State fMRI in Pre-Adolescents with ADHD

**DOI:** 10.1101/2023.08.17.23294136

**Authors:** Ru Zhang, Stuart B. Murray, Christina J. Duval, Danny J.J. Wang, Kay Jann

## Abstract

Attention deficit hyperactivity disorder (ADHD) has been characterized by impairments among distributed functional brain networks, e.g., the frontoparietal network (FPN), default mode network (DMN), and reward and motivation-related circuits (RMN). In the current study, we evaluated the complexity and functional connectivity (FC) of resting state fMRI (rsfMRI) in pre-adolescents with ADHD for pathology-relevant networks.

We leveraged data from the Adolescent Brain and Cognitive Development (ABCD) Study. The final study sample included 63 children with ADHD and 92 healthy control children matched on age, sex, and pubertal development status. For selected regions in relevant networks, ANCOVA compared multiscale entropy (MSE) and FC between the groups. Finally, differences in the association between MSE and FC were evaluated.

We found significantly reduced MSE along with increased FC within the FPN of pre-adolescents with ADHD compared to matched healthy controls. Significant partial correlations between MSE and FC emerged in fewer regions in the participants with ADHD than in the controls.

The observation of reduced entropy is consistent with existing literature using rsfMRI and other neuroimaging modalities. The current findings of complexity and FC in ADHD support hypotheses of altered function of inhibitory control networks in ADHD.

## 1. Introduction

Attention deficit hyperactivity disorder (ADHD) is characterized by a persistent pattern of inattention and/or hyperactivity and impulsivity that causes impairment in functioning and development. ADHD occurs worldwide and is estimated to affect 5% of children and approximately 2.5% of adults (Polanczyk, de Lima, Horta, Biederman, & Rohde, 2007). Although numerous studies have been conducted on ADHD, the psychopathology of the disorder is still not fully understood. In recent years, ADHD has been increasingly viewed as impairments among distributed functional networks or circuits (Friedman & Rapoport, 2015; Paloyelis, Mehta, Kuntsi, & Asherson, 2007). Neuroimaging has been used to identify networks, which consist of brain regions that show synchronized, correlated fMRI signal dynamics (Biswal, Yetkin, Haughton, & Hyde, 1995; Fox, Zhang, Snyder, & Raichle, 2009). These functionally connected networks (FCNs) are believed to reflect distinct mental states or processes (Buckner & Krienen, 2013; Shirer, Ryali, Rykhlevskaia, Menon, & Greicius, 2012). Aberrant communication between or within FCNs may reflect deficits in cognitive and affective functioning (Kessler, Angstadt, Welsh, & Sripada, 2014; Zang et al., 2007).

In ADHD, altered functional connectivity has been reported in the default mode network (DMN), frontoparietal network (FPN), and reward and motivation-related circuits (RMN; Castellanos & Aoki, 2016; Posner, Park, & Wang, 2014). Importantly, FCN findings in ADHD are inconsistent across different studies and age groups. Three recent meta-analyses summarized the rsfMRI functional connectivity findings of ADHD vs. healthy controls based on seed-based analyses (Cortese, Aoki, Itahashi, Castellanos, & Eickhoff, 2021; Gao et al., 2019; Sutcubasi et al., 2020). Gao et al. (2019) found ADHD was associated with hyperconnectivity between the seeds in DMN and the left superior temporal gyrus and right supramarginal gyrus extending to the right angular gyrus. ADHD also showed hypoconnectivity between the seeds in DMN to the left middle frontal gyrus and subcallosal cingulate cortex. With respect to the seeds in FPN, Gao et al. (2019) reported hyperconnectivity to the left orbital frontal cortex, caudate, putamen, and insula and hypoconnectivity to the precentral gyri among those with ADHD. For the seeds in RMN, hyperconnectivity to the dorsolateral prefrontal cortex was observed in ADHD. Sutcubasi et al. (2020) reported somewhat similar results in DMN and FPN but no agreement for RMN. In terms of the seeds in DMN, they found reduced connectivity to the dorsal posterior cingulate cortex but elevated connectivity to the dorsomedial prefrontal cortex in ADHD, while for the seeds in FPN, ADHD also showed elevated connectivity to the anterior prefrontal cortex. Cortese et al. (2021) did find no significant spatial convergence of ADHD-related hyperconnectivity or hypoconnectivity across studies. However, these inconclusive patterns of hyper and hypoconnectivity in ADHD also persist when focusing on children and adolescents only (Gao et al., 2019; Sutcubasi et al., 2020) but suggesting a more widespread connectivity alteration as compared to adult ADHD groups that could potentially be attributed to neurodevelopmental changes or delays in functional connectivity from childhood to adulthood (Fair et al., 2009).

Alongside functional connectivity analyses that assess the signal coherence between nodes, nonlinear analyses of neural signals from fMRI that characterize the signal complexity within a node have been proposed as measures for information processing capacity of brain areas and networks (McDonough & Nashiro, 2014; Smith, Yan, & Wang, 2014; D. J. J. Wang et al., 2018; Z. Wang, Li, Childress, & Detre, 2014), or indices of pathological brain function (Fernández, Gómez, Hornero, & López-Ibor, 2013; Sun et al., 2020; Takahashi, 2013). Sample entropy (SampEn; Richman & Moorman, 2000) has attracted considerable attention in complexity analysis because of its simplicity and reduced dependency on time series length compared to other forms of entropy such as approximation entropy (Pincus, 1991). SampEn measures the randomness and predictability of a stochastic process and generally increases with greater complexity. Multiscale entropy (MSE) was subsequently introduced to differentiate complex processes from random fluctuations more accurately, by calculating SampEn of a signal at multiple coarse-grained time scales (Costa, Goldberger, & Peng, 2002). Because MSE is evaluated across different time scales, it is also capable to identify frequency-dependent neuropathophysiological processes in different brain regions. Two studies investigated the complexity of fMRI data in adults with ADHD (Guan et al., 2023; Sokunbi et al., 2013) and both reported reduced complexity in the resting state in the frontal cortical areas, but no study as of yet has investigated fMRI complexity differences in children or pre-adolescents when ADHD is initially diagnosed. Additionally, no study has assessed the relation between functional connectivity and MSE (McDonough & Nashiro, 2014; D. J. J. Wang et al., 2018) in pre-adolescents with ADHD.

In the current study, we aimed to address this critical gap in the extant literature and evaluated the functional connectivity and complexity of rsfMRI in pre-adolescents with ADHD for the three main networks commonly associated with ADHD: FPN, DMN, and RMN. We combined MSE complexity analysis with functional connectivity analysis to elucidate the functional alterations present in ADHD on network nodes, at node-to-node edges, and on nodal complexity and node-to-node connectivity interaction levels. According to the literature, we hypothesized that compared to the healthy controls, pre-adolescents with ADHD had lower MSE in the FPN and altered functional connectivity in the FPN, DMN, and RMN. Due to the lack of previous studies, we did not postulate a hypothesis in terms of the difference between ADHD and control groups on the association between MSE and functional connectivity.

## 2. Methods

### 2.1 Participants

We leveraged the baseline demographic, clinical, T1 structural, and rsfMRI data from the Adolescent Brain and Cognitive Development (ABCD) Study (Casey et al., 2018). The ABCD study is the largest pediatric brain imaging study in the United States, involving 21 research sites across the country. ADHD diagnoses were confirmed according to DSM-5 criteria (American Psychiatric Association. & American Psychiatric Association. DSM-5 Task Force., 2013), by the Kiddie-Schedule for Affective Disorders and Schizophrenia (KSADS)-parent report. Subjects with ADHD and any other psychiatric comorbidity were excluded from the current study. The final study sample included 63 subjects with ADHD (age = 9.91 ± 0.640 months; pubertal development status [PDS] = 1.665 ± 0.542; 25 female/38 male) and 92 controls (age = 10.009 ± 0.651 months; PDS = 1.687 ± 0.522; 29 female/63 male) matched in terms of age (*t* = 0.859, *p* = 0.392), sex (χ^2^ = 0.767, *p* = 0.381), and PDS (*t* = 0.255 *p* = 0.799; see Table 1).

**Table 1.** An overview of sample characteristics delineated by group.

|  | ADHD group (*N* = 63) |  | Control group (*N* = 92) |  | Statistic | *p* value |
| --- | --- | --- | --- | --- | --- | --- |
| Sex (f/m) | 25/38 | | 29/63 | | $\chi^2 = 0.767$ | 0.381 |
|  | Mean | Standard deviation | Mean | Standard deviation |  |  |
| Age (years) | 9.910 | 0.640 | 10.009 | 0.651 | $t = 0.859$ | 0.392 |
| Pubertal development status | 1.665 | 0.542 | 1.687 | 0.522 | $t = 0.255$ | 0.799 |
| ADHD score | 5.412 | 2.152 | 1.228 | 1.570 | $t = -13.995$ | < 0.001 |

Key to Table 1: Pubertal development status was measured by the self-reported pubertal development scale (Petersen, Crockett, Richards, & Boxer, 1988). The ADHD score was measured by the parent-reported ADHD CBCL-DSM5 scale (Achenback & Rescorla, 2001).

### 2.2 Imaging Acquisition, Preprocessing, and Denoising

All T1-weighted scans were acquired with voxel resolution = 1mm^3^, 256□×□256 matrix, flip angle = 8°, and 2x parallel imaging. Other scan parameters slightly varied by scanner platform, i.e., Siemens Prisma, Philips, or GE 3T scanner^16^. Each participant had 3-4 eyes-open (passive crosshair viewing) rsfMRI scans, each of which was approximately 5-minutes in duration. All rsfMRI scans were collected using a gradient-echo EPI sequence of 383 volumes in total (voxel resolution = 2.4 × 2.4 × 2.4□mm^3^, 60□slices, 90□×□90 matrix, FOV = 216□×□216, TR = 800□ms, TE = 30□ms, flip angle = 52°, 6-factor multiband acceleration). Head motion was monitored during scan acquisition using real-time procedures to adjust scanning procedures as necessary.

For the current study, we used rsfMRI run1 and run2. Spatial and temporal image preprocessing was performed in the CONN toolbox (Conn: fMRI functional connectivity toolbox). Default parameter setup was used including motion-realignment, coregistration and normalization to MNI152 template space and smoothing with a 6mm FWHM Gaussian kernel. In the denoising process, white matter, CSF, and motion realignment parameters were included for temporal preprocessing and denoising of rsfMRI data. While motion-scrubbing was included in the preprocessing for functional connectivity analysis, this step was excluded for complexity analysis.

### 2.3 Data Analysis

#### 2.3.1 Complexity Analysis

Voxel-wise multiscale entropy (MSE) was computed using the in-house developed LOFT Complexity Toolbox (github.com/kayjann/complexity) for each preprocessed and denoised rsfMRI sequence for each participant. SampEn is defined as the natural logarithm of the conditional probability that a pattern length of *m* points will repeat itself, excluding self-matches, for *m* + 1 points within a tolerance of *r* in a time series of length *N* (Richman & Moorman, 2000). Multiple “coarse-grained” time series of lengths *N*/1, *N*/2,…, and *N*/*a* were formed by averaging consecutive data points of increasing length. SampEn for each coarse-grained time series is calculated and MSE is the average of SampEn over all the temporal scales.

In the current MSE calculation, we set the pattern length *m* = 2, the sensitivity threshold *r* = 0.3, and the number of temporal scales *a* = 15. MSE was computed for each voxel for each rsfMRI time series for each participant. The MSE map was then obtained by averaging MSE over the two rsfMRI runs for each voxel for each participant. Finally, mean MSE within each region-of-interest (ROI) of the three networks was calculated for each participant: ROIs were selected in the anterior cingulate gyrus for both FPN and DMN, middle frontal gyrus and superior frontal gyrus for FPN, supramarginal gyrus, medial frontal cortex, and posterior cingulate gyrus for DMN, and frontal orbital cortex, caudate, and putamen for RMN based on the Harvard-Oxford atlas (Desikan et al., 2006).

A factorial 2 (Group: ADHD, Control) by 2 (Sex: Male, Female) analysis of covariance (ANCOVA) on the mean MSE in each ROI was conducted with PDS (self-reported) and scan-site as covariates. We were only interested in the main effects of Group here and in this study’s other ANCOVA analyses. To reduce skewness and kurtosis (Bliss, Greenwood, & White, 1956), PDS scores were Rankit-transformed and then z-scored. These values were used as the continuous covariates in all the ANCOVAs in the current study. A Benjamini-Hochberg correction (Benjamini & Hochberg, 1995) was performed for each network individually. The same ANCOVA was repeated for SampEn at each temporal scale.

In order to evaluate the association between complexity and ADHD behavior symptoms, the partial correlation between MSE and CBCL DSM5 ADHD scores controlling for sex, PDS z-score, and scan-site was computed for the whole sample. We also computed the partial correlation between SampEn at each temporal scale and the CBCL DSM5 ADHD scores controlling for the same covariates for the whole sample. Note that the CBCL DSM5 ADHD scores were Rankit-transformed and then z-scored in this partial correlation analysis.

#### 2.3.2 Functional Connectivity Analysis

After data preprocessing and denoising, the first-level analyses on functional connectivity were conducted for each participant across both runs using the CONN toolbox default setup. Time series were averaged over voxels in each ROI, correlated between each pair of ROIs using Pearson’s correlation analysis, and then Fisher’s Z-transformed. The same ANCOVA including PDS z-score and scan-site as covariates was then performed for functional connectivity (Fisher’s Z-transformed) using a seed-to-seed analysis. A Benjamini-Hochberg correction (Benjamini & Hochberg, 1995) was performed on the resultant main effects of Group for each network.

In order to evaluate the association between functional connectivity and ADHD behavior symptoms, the partial correlation between functional connectivity and CBCL DSM5 ADHD scores controlling for sex, PDS z-score, and scan-site was computed for the whole sample. Note again that the CBCL DSM5 ADHD scores were Rankit-transformed and then z-scored in this partial correlation analysis.

#### 2.3.3 Analysis of the Association between Complexity and Functional Connectivity

To investigate the association between complexity and functional connectivity, we computed the partial correlation (i.e., Fisher’s *z* score) between functional connectivity (between each seed ROI and each other ROI) and MSE in each seed ROI for each group by controlling for sex, PDS z-score, and scan-site. A group comparison (ADHD vs. controls) was conducted on the partial correlations for each (seed-MSE, seed-to-seed connectivity) linkage. The same analysis was repeated for the partial correlation between SampEn at each temporal scale and functional connectivity.

#### 2.3.4 Follow-Up Analyses

In order to investigate the potential medication effects, a factorial 2 (Medication: medicated ADHD, non-medicated ADHD) by 2 (Sex: Male, Female) ANCOVA on the mean MSE in each ROI was conducted with PDS z-score and scan-site as covariates. The same ANCOVA was repeated for SampEn at each temporal scale and for functional connectivity at each seed-to-seed edge.

## 3. Results

### 3.1 Clinical Data

There were no group differences in age (*t* = -0.859, *p* = 0.392), sex (χ^2^ = 0.767, *p* = 0.381), or PDS (*t* = -0.255, *p =* 0.799; see Table 1). As expected, participants with ADHD scored significantly higher than the controls on the CBCL DSM5 ADHD score (*t* = 13.995, *p <* 0.001; see Table 1). There was no significant group difference in terms of the framewise displacement (*p* > 0.05 for both runs; Power et al., 2014).

### 3.2 Complexity Analysis

The average MSE maps over the two rsfMRI runs for the ADHD and control groups are displayed in Figures 1(a) and (b). Qualitatively, it can be seen that complexity in the grey matter was higher than in the white matter. Moreover, the ADHD group appeared to have lower MSE than the controls, specifically in the frontal cortex. The ANCOVA revealed significant main effects of Group in multiple ROIs, i.e., the anterior cingulate gyrus (FPN & RMN; *F* = 4.417, *p* < 0.05, 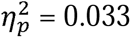), bilateral middle frontal gyrus (FPN; *F* = 5.491 & 5.557, *p* < 0.05, 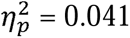 & 0.041, respectively), bilateral superior frontal gyrus (FPN; *F* = 4.423 & 4.831, *p* < 0.05, 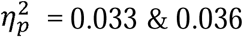 & 0.036, respectively), and frontal medial cortex (RMN; *F* = 4.626, *p* < 0.05, 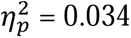). The Benjamini-Hochberg correction was performed for each network separately (false discovery rate = 0.05) and all the ROIs in FPN survived the correction. Post-hoc *t-*tests indicated that the participants with ADHD had lower MSE values than the controls in all the ROIs in FPN (MSE of ADHD vs. control = -0.078 to -0.042; *t* = -1.768 to -0.715; see Figure 1(c) and Table 2).

**Figure 1.**
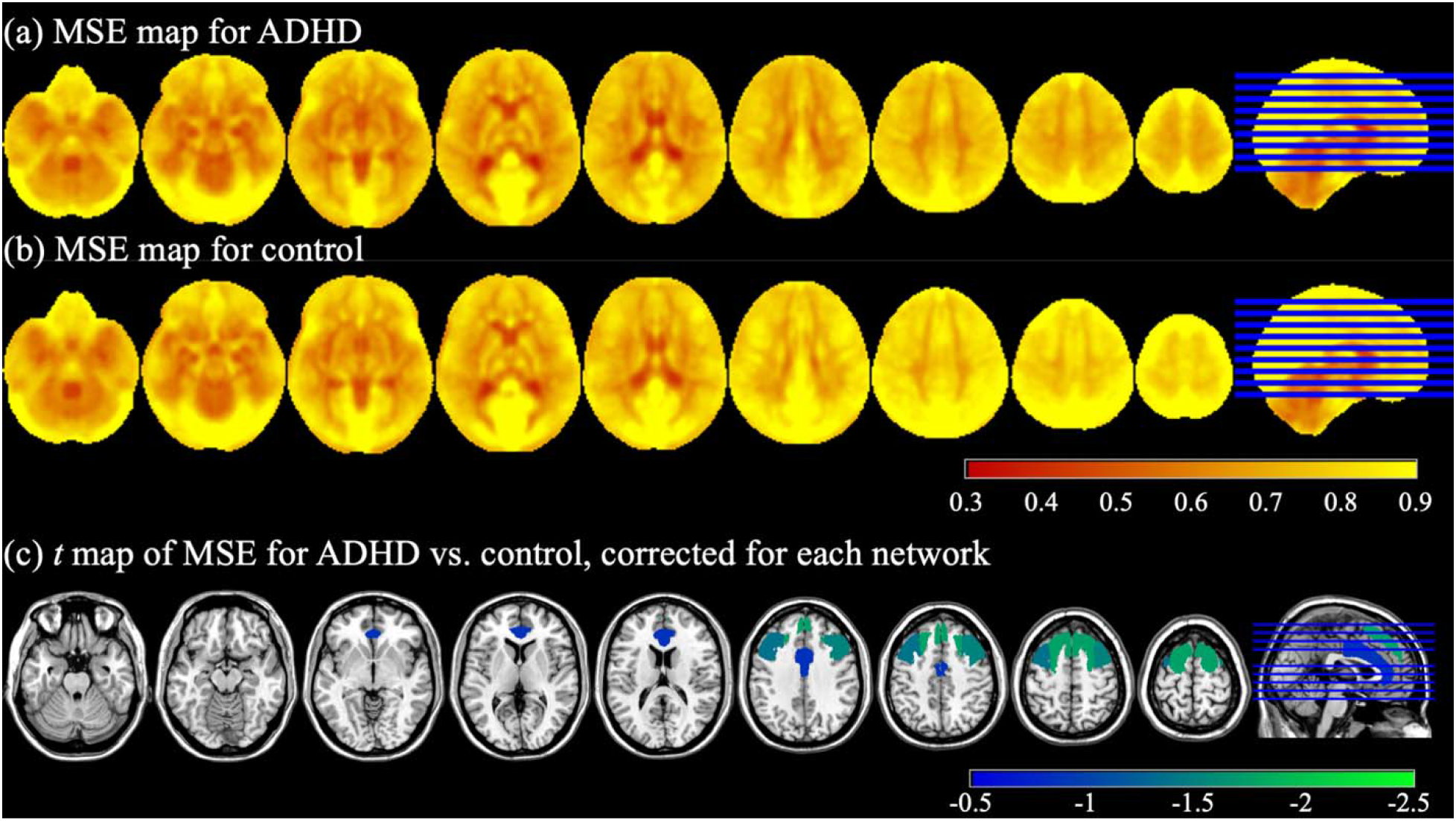
(a) MSE map for the ADHD group. (b) MSE map for the control group. (c) *t* value map of MSE for ADHD vs. control. Only significant main effects of Group from ANCOVA were displayed, Benjamini-Hochberg corrected for each network (*p* < 0.05, false discovery rate = 0.05).

**Table 2.**
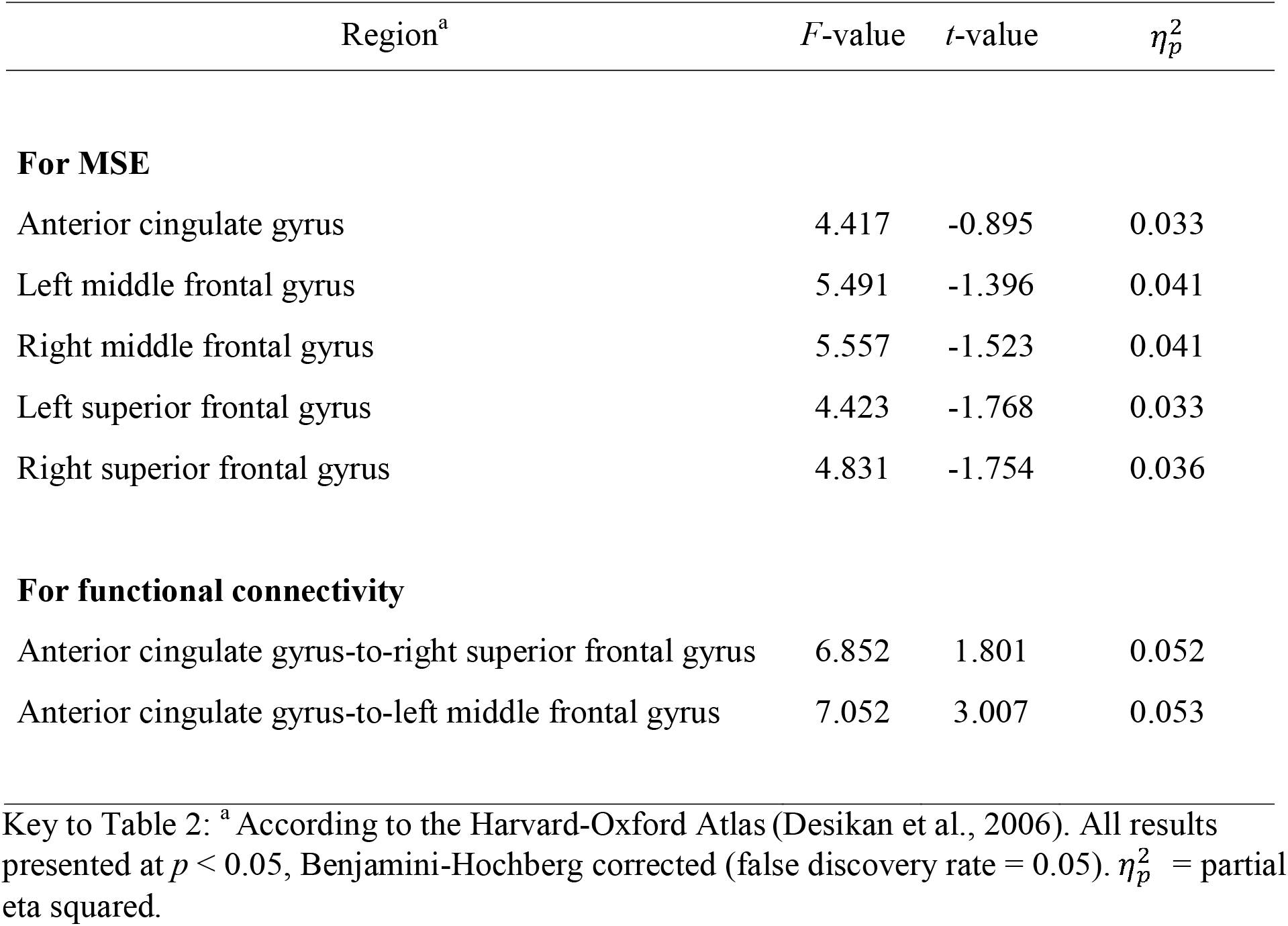
Brain regions displaying significant main effects of Group, obtained from the ANCOVAs for MSE and functional connectivity.

| Region<sup>a</sup> | *F*-value | *t*-value | $\eta_p^2$ |
| --- | --- | --- | --- |
| **For MSE** |  |  |  |
| Anterior cingulate gyrus | 4.417 | -0.895 | 0.033 |
| Left middle frontal gyrus | 5.491 | -1.396 | 0.041 |
| Right middle frontal gyrus | 5.557 | -1.523 | 0.041 |
| Left superior frontal gyrus | 4.423 | -1.768 | 0.033 |
| Right superior frontal gyrus | 4.831 | -1.754 | 0.036 |
| **For functional connectivity** |  |  |  |
| Anterior cingulate gyrus-to-right superior frontal gyrus | 6.852 | 1.801 | 0.052 |
| Anterior cingulate gyrus-to-left middle frontal gyrus | 7.052 | 3.007 | 0.053 |

Key to Table 2: <sup>a</sup> According to the Harvard-Oxford Atlas (Desikan et al., 2006). All results presented at $p < 0.05$ , Benjamini-Hochberg corrected (false discovery rate = 0.05). $\eta_p^2$ = partial eta squared.

Figure S1 in the supplementary material displays the *t* value of ADHD vs. control for mean MSE and SampEn at each temporal scale in each ROI which showed a significant main effect of Group (*p* < 0.05, uncorrected). It reveals that at scale 1 (frequency = 1.25Hz), every ROI showed a significant main effect of Group except the left anterior supramarginal gyrus. As the scale number increased, some significant main effects of Group disappeared although all the ROIs in FPN remained till scale = 9 (frequency = 0.111Hz). At scale = 15 (frequency = 0.083Hz), there was no significant difference between the ADHD group and the control group in terms of SampEn (*p* > 0.05, uncorrected).

We further tested whether there was an association between MSE and the CBCL DSM5 ADHD score but the partial correlation analysis for the whole sample taking into account sex, PDS z-score, and scan-site did not reach statistical significance in any ROI (*p* > 0.05). We also computed the same partial correlation between SampEn and the CBCL DSM5 ADHD score at each temporal scale and none was significant (*p* > 0.05).

### 3.3 Functional Connectivity Analysis

Figures 2(a) and (b) display the significant functional connectivity over the two rsfMRI runs for the ADHD group and the control group, respectively, Benjamini-Hochberg corrected for each network (*p* < 0.05, false discovery rate = 0.05). It can be seen that most ROIs were strongly connected within each network. The ANCOVA revealed significant main effects of Group at two edges in FPN after Benjamini-Hochberg correction (false discovery rate = 0.05), i.e., anterior cingulate gyrus-to-right superior frontal gyrus (FPN; *F* = 6.852, *p* < 0.05, 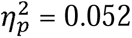) and anterior cingulate gyrus-to-left middle frontal gyrus (FPN; *F* = 7.052, *p* < 0.05, 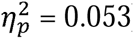). Post-hoc *t-*tests indicated that the participants with ADHD had stronger functional connectivity than the controls at these edges (functional connectivity of ADHD vs. control = 0.097 & 0.094; *t* = 1.801 & 3.007, respectively; see Figure 2 (c) and Table 2). For the uncorrected results, interested readers shall refer to Figure S2 in the supplementary material for the functional connectivity map for each group and the *t*-value map for ADHD vs. control and Table S1 in the supplementary material for the statistical summary of the main effects of Group (*p* < 0.05, uncorrected).

**Figure 2.**
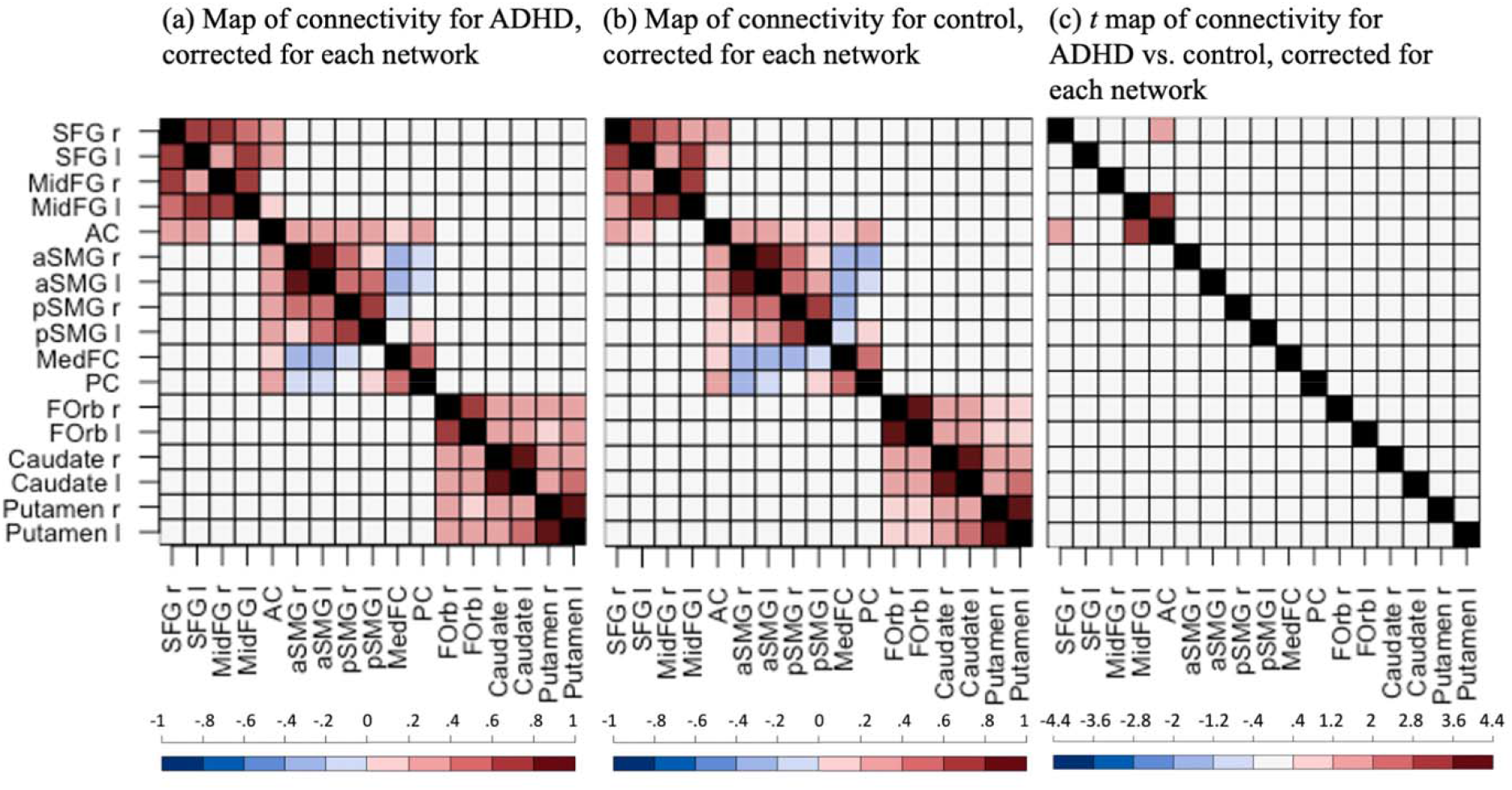
Functional connectivity maps (Fisher’s z) for (a) the ADHD group and (b) the control group, and (c) the *t*-value map for the ADHD group vs. the control group. Only significant edges are displayed in blue or red colors, Benjamini-Hochberg corrected for each network (*p* < 0.05, false discovery rate = 0.05). For (a) and (b), blue/red indicates *p* < 0.05 for the *t*-test against 0. For (c), blue/red indicates *p* < 0.05 for the main effect of Group. The black indicates data was not available. SFG: superior frontal gyrus; MidFG: middle frontal gyrus; AC: anterior cingulate gyrus; aSMG: anterior supramarginal gyrus; pSMG: posterior supramarginal gyrus; MedFC: frontal medial cortex; PC: posterior cingulate gyrus; FOrb: frontal orbital cortex; r: right; l: left.

The partial correlations between functional connectivity and the CBCL DSM5 ADHD score controlling for sex, PDS z-score, and scan-site were significant and positive at the edges of the left posterior supramarginal gyrus-to-the right caudate (*r* = 0.161, *p* = 0.045), the left posterior supramarginal-to-the left caudate (*r* = 0.160, *p* = 0.046), and the right frontal orbital cortex-to-the left caudate (*r* = 0.217, *p* = 0.007). However, after Benjamini-Hochberg correction, only the edge of the right frontal orbital cortex-to-the left caudate, part of the RMN, survived.

### 3.4 Analysis of the Association between Complexity and Functional Connectivity

Figures 3(a) and (b) display the significant partial correlation (Fisher’s z) between MSE (in a specific seed) and functional connectivity (at the edge of this seed to another ROI) for each group after controlling sex, PDS z-score, and scan-site, Benjamini-Hochberg corrected for each network (*p* < 0.05, false discovery rate = 0.05). Compared to the controls, the ADHD group appeared to have fewer (seed-MSE, seed-to-seed connectivity) linkages that were significantly different from zero. However, Benjamini-Hochberg corrected results (corrected for each network; false discovery rate = 0.05) indicated no significant between-group difference for any linkage (see Figure 3(c)). Please refer to Table S2 in the supplementary material for the summary of the uncorrected results.

**Figure 3.**
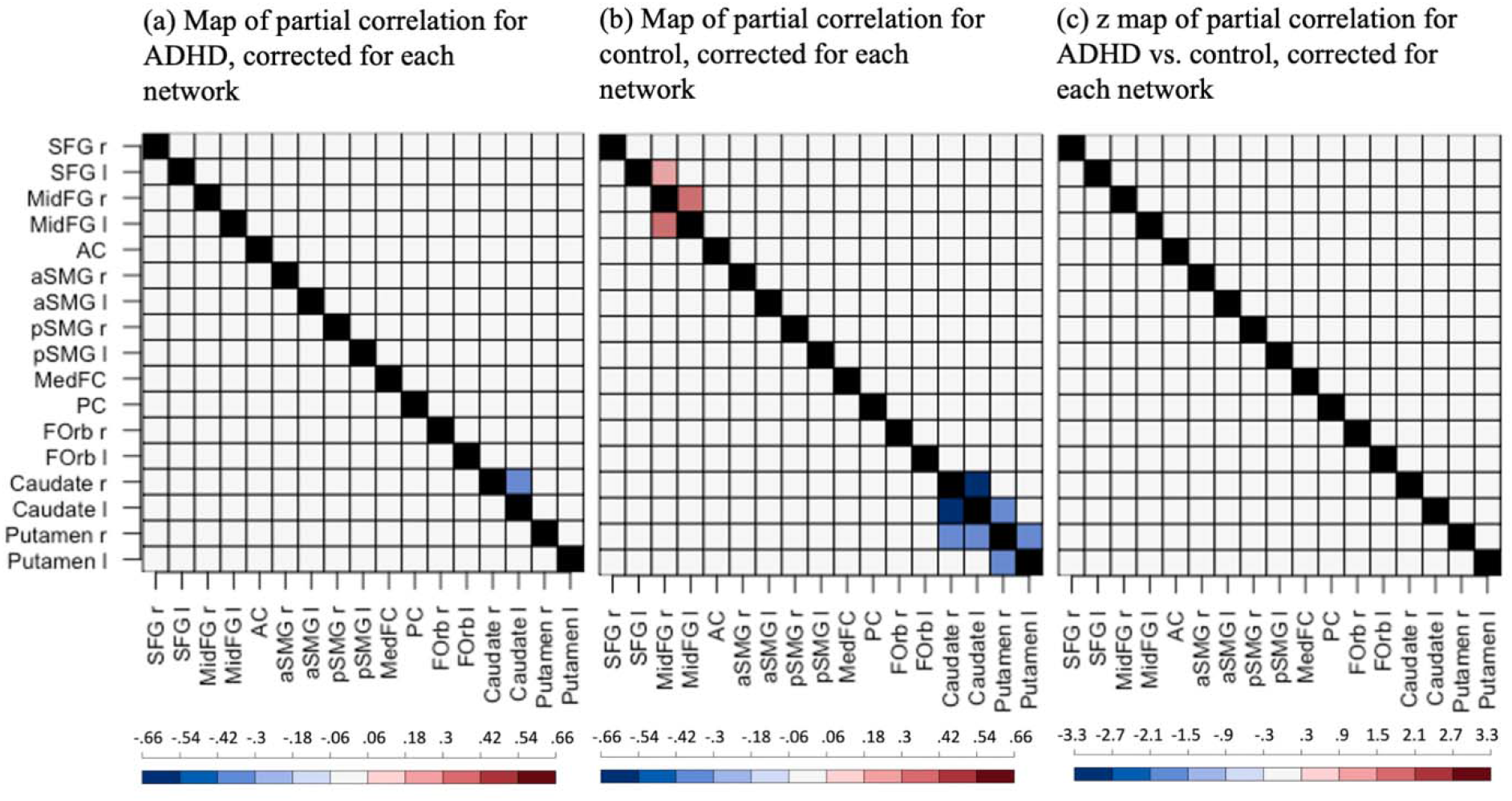
Maps of partial correlations (Fisher’s z) between MSE and functional connectivity for (a) the ADHD group and (b) the control group, and (c) the z value map of partial correlations for ADHD vs. control. Only significant (seed-MSE, seed-to-seed connectivity) linkages are displayed in blue or red colors, Benjamini-Hochberg corrected for each network (*p* < 0.05, false discovery rate = 0.05). For (a) and (b), blue/red indicates *p* < 0.05 for the *t*-test against 0. For (c), blue/red indicates *p* < 0.05 for the statistical comparison of partial correlations in the ADHD and control groups. The black indicates data was not available. SFG: superior frontal gyrus; MidFG: middle frontal gyrus; AC: anterior cingulate gyrus; aSMG: anterior supramarginal gyrus; pSMG: posterior supramarginal gyrus; MedFC: frontal medial cortex; PC: posterior cingulate gyrus; FOrb: frontal orbital cortex; r: right; l: left.

Figure S3 in the supplementary material displays the significant partial correlations (Fisher’s z) between MSE or SampEn at each temporal scale and functional connectivity for each group (*p* < 0.05, uncorrected; Figures S3(a) and (b)) and the z values of partial correlations for ADHD vs. control (only significant linkages are shown, *p* < 0.05, uncorrected; Figure S3(c)). Across all the temporal scales, the ADHD group appeared to have fewer (seed-MSE, seed-to-seed connectivity) linkages within networks and between networks, significantly different from zero than the controls. Moreover, ADHD showed significantly lower partial correlations between SampEn and functional connectivity relative to the controls within DMN at each temporal scale except scale = 1 (*p* < 0.05, uncorrected), specifically in linkages of (frontal medial cortex, frontal medial cortex-to-right anterior supramarginal gyrus), (frontal medial cortex, frontal medial cortex-to-left anterior supramarginal gyrus), (right anterior supramarginal gyrus, right anterior supramarginal gyrus-to-medial frontal cortex), and (left anterior supramarginal gyrus, left anterior supramarginal gyrus-to-medial frontal cortex). ADHD showed significantly higher partial correlations relative to the controls within RMN (*p* < 0.05, uncorrected): The linkage of (right putamen, right putamen-to-right caudate) was significant at each temporal scale; Some significant within-RMN linkages emerged at the higher scale number (scale > 10), i.e., (left putamen, left putamen-to-right caudate), (right caudate, right caudate-to-right putamen), and (right caudate, right caudate-to-left putamen). ADHD showed a significantly higher partial correlation relative to the controls within FPN only at scale = 1 in the linkage of (right superior frontal gyrus, right superior frontal gyrus-to-anterior cingulate gyrus) (*p* < 0.05, uncorrected). In terms of cross-network linkages, we observed ADHD had a lower partial correlation between SampEn and functional connectivity than the controls in the linkages of (frontal medial cortex, frontal medial cortex-to-right putamen) and (frontal medial cortex, frontal medial cortex-to-left putamen) at almost every temporal scale up to scale = 13 (*p* < 0.05, uncorrected). We also observed ADHD had a higher partial correlation than the controls in the linkage of (right putamen, right putamen-to-right middle frontal gyrus) at scale = 11 & 12 (*p* < 0.05, uncorrected).

### 3.5 Follow-Up Analyses

In order to investigate potential medication-related effects, a factorial 2 (Medication: medicated ADHD, non-medicated ADHD) by 2 (Sex: Male, Female) ANCOVA on the mean MSE in each ROI was conducted with PDS z-score and scan-site as covariates. Please refer to Table S3 in the supplementary material for the list of prescriptions. The ADHD participants who consumed at least one prescription were categorized as medicated ADHD. The result indicated a main effect of Medication in the left anterior supramarginal gyrus (DMN; *F* = 4.563, *p* < 0.05, 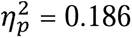). However, it did not survive the networkwise Benjamini-Hochberg correction (false discovery rate = 0.05). Therefore, we concluded there was no significant difference in MSE between the medicated ADHD participants and the non-medicated ADHD participants in any ROI. The same ANCOVA was replicated for SampEn at each temporal scale. Main effects were found in multiple regions at multiple temporal scales (*p* < 0.05, uncorrected). More details can be accessed from the supplementary material. However, none of them survived the Benjamini-Hochberg correction (false discovery rate = 0.05).

The same ANCOVA was replicated for functional connectivity at each seed-to-seed edge. It revealed significant main effects of Medication at two edges, i.e., left middle frontal gyrus-to-right frontal orbital cortex (FPN-to-RMN; *F* = 4.509, *p* < 0.05, 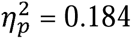) and medial frontal gyrus-to-posterior cingulate gyrus (DMN-to-DMN; *F* = 4.574, *p* < 0.05, 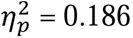). However, none of them survived the Benjamini-Hochberg correction (false discovery rate = 0.05).

## 4. Discussion

In this study, we report two main observations: (1) reduced rsfMRI complexity in areas of the FPN in pre-adolescents with ADHD as compared to the matched controls, and (2) increased functional connectivity between anterior portions of the FPN in the ADHD group relative to the control group. We did not observe a significant group difference in terms of the association between complexity and functional connectivity in any interested linkage.

The FPN has been hypothesized to play a role in instantiating and flexibly modulating cognitive control (Marek & Dosenbach, 2018). Cognitive control is commonly compromised in many forms of psychopathology, many of which emerge during childhood and adolescence, while the control function is refined throughout development. Emerging literature has suggested an important role of the FPN in the pathophysiology of ADHD. Previous structural MRI studies consistently reported ADHD was associated with abnormal morphometry (Albajara Sáenz, Villemonteix, & Massat, 2019; Frodl & Skokauskas, 2012) and developmental trajectories (M. Shaw et al., 2012; P. Shaw et al., 2007) in the prefrontal and cingulate structures. Both qualitative review (Rubia, 2011) and meta-analyses (Cortese et al., 2012; Hart, Radua, Nakao, Mataix-Cols, & Rubia, 2013) on task-fMRI reported hypoactivation in the regions of FPN in ADHD across different tasks. rsfMRI meta-analyses also suggested functional alternation of FPN in ADHD (Cortese et al., 2021; Gao et al., 2019; Sutcubasi et al., 2020). Specifically, Gao et al. (2019) reported children/adolescents with ADHD demonstrate hyperconnectivity between the seeds in the FPN and the dorsal anterior cingulate cortex, a finding that our results corroborate. In addition to altered functional connectivity in the FPN, the current study revealed significantly different complexity in pre-adolescents with ADHD relative to the controls thus extending existing literature and providing additional evidence for the hypothesis of altered cognitive control in ADHD.

The observation of significant reductions in MSE in rsfMRI time series in pre-adolescents with ADHD was consistent with the findings in adults with ADHD using the same modality (Guan et al., 2023; Sokunbi et al., 2013). Guan et al. (2023) observed significant decreases of SampEn in multiple ROIs in every investigated brain network under every atlas in subjects with ADHD compared to controls. They also observed reduced SampEn of the gray matter at every temporal scale under every atlas in ADHD. Sokunbi et al. (2013) reported significant reductions of SampEn in adults with ADHD relative to the controls in the superior frontal and anterior cingulate gyrus which was replicated for pre-adolescents with ADHD in the current study (Figure 1(C)). Our results aligned with existing literature in resting-state using other neuroimaging modalities such as EEG and MEG in which complexity was found lower in the participants with ADHD than in the healthy controls (Angulo-Ruiz, Muñoz, Rodríguez-Martínez, Cabello-Navarro, & Gómez, 2022; Fernández et al., 2009; Gu, Liu, & Woltering, 2022; Gómez et al., 2013; Hu et al., 2021; Rezaeezadeh, Shamekhi, & Shamsi, 2020).

While multiple previous studies support the conceptualization of ADHD as a dysconnectivity syndrome, findings relating to functional connectivity under rsfMRI have been highly heterogeneous in terms of the affected networks and network subcomponents, as well as the direction of effects. The current study observed *enhanced* connectivity within FPN, i.e., the right superior frontal gyrus and left middle frontal gyrus using the anterior cingulate gyrus as the seed in pre-adolescents with ADHD (Figure 2). This result was consistent with previous literature that reported enhanced connectivity between the anterior cingulate gyrus and middle frontal gyrus in children/adolescents with ADHD compared to healthy controls (Kumar, Arya, & Agarwal, 2021; Lin, Kessler, Tseng, & Gau, 2021; Liu et al., 2023) except for one study reporting decreased connectivity (Posner et al., 2013). Interestingly this increased functional connectivity is predominantly between anterior nodes of the network and one might speculate that this represents a developmental stage of the network of increased short-range connectivity before long-range distributed network connections are established (Fair et al., 2009).

To date, several studies have investigated the differences between psychiatric and non-psychiatric cohorts in terms of complexity or/and functional connectivity using functional MRI time series. However, while an association between MSE and functional connectivity in healthy cohorts has been previously reported (McDonough & Nashiro, 2014; D. J. J. Wang et al., 2018), as far as we have known, the current study is the first effort to conduct a comparison on the association between complexity and functional connectivity between healthy and diseased groups. While we found significant positive relationships between MSE and functional connectivity in the FPN and negative associations in the RMN in the healthy control group, such relationships were almost entirely absent in the group with ADHD (Figure 3). However, the direct statistical comparison did not reveal a significant group difference in the relation between MSE and functional connectivity between participants with ADHD and the controls. Future research in ADHD or other neuropsychiatric disorders following this approach might reveal disease and network-specific alterations in the association between nodal complexity and node-to-node connectivity, two complementary but interrelated network properties.

Several caveats should be considered with respect to the current results. First, many ADHD participants were prescribed ADHD medications. In order to evaluate the effect of prescription, a factorial 2 (Medication: medicated ADHD, non-medicated ADHD) by 2 (Sex: Male, Female) ANCOVA on the mean MSE, SampEn at each temporal scale, and functional connectivity in each ROI was conducted with PDS z-score and scan-site as covariates. The result after correction indicated no significant main effects of Medication for MSE, SampEn at each temporal scale, and functional connectivity in any ROI. Second, it is worth mentioning that the patients included in the current work had no previous or current psychiatric diagnoses other than ADHD. Many previous complexity studies in ADHD indeed examined ADHD with potential various comorbidities such as oppositional defiant disorder/conduct disorder that might confound the results. The current results removed this concern and further confirmed the finding of reduced complexity. Moreover, the lack of a unified understanding of functional connectivity in ADHD across literature may be due in part to different rates of comorbid conditions in those studies. The current finding of enhanced functional connectivity in ADHD relative to the controls in the FPN might potentially contribute to clarifying the mystery and provide a greater understanding of this field. Third, due to the limitation of networkwise Benjamini-Hochberg correction, the group difference in terms of cross-network functional connectivity and cross-network partial correlation between MSE and functional connectivity was absent. However, we showed significant cross-network group differences in Tables S1 and S2 and Figures S2 and S3 in the supplementary material. The reader should keep in mind that those effects were based on uncorrected statistics.

In conclusion, our analyses support the feasibility and utility of temporal signal entropy in investigating the changes in complexity of rsfMRI signals in pre-adolescents with ADHD when compared to healthy controls. We found reduced complexity in the anterior cingulate gyrus, bilateral middle frontal gyrus, and bilateral superior frontal gyrus in the FPN and greater functional connectivity in the right superior frontal gyrus and left middle frontal gyrus using the anterior cingulate gyrus as the seed in pre-adolescents with ADHD. We suggest that combining analyses of complexity and functional connectivity is a useful and easily obtainable measure to reveal changes in ADHD brain dynamics.

## Data Availability

All data produced are available online at https://abcdstudy.org

## Acknowledgments

Funding: This study was funded by NIA R01AG066711 (KJ/DJW) and SBM is supported by the Della Martin Endowed Professorship, and the National Institute of Mental Health (K23MH115184).

Additional Information: The ABCD Study was supported by the National Institutes of Health and additional federal partners under award numbers U01DA041022, U01DA041025, U01DA041028, U01DA041048, U01DA041089, U01DA041093, U01DA041106, U01DA041117, U01DA041120, U01DA041134, U01DA041148, U01DA041156, U01DA041174, U24DA041123, and U24DA041147. A full list of supporters is available at https://abcdstudy.org/nihcollaborators. A listing of participating sites and a complete listing of the study investigators can be found at https://abcdstudy.org/principal-investigators.html. ABCD consortium investigators designed and implemented the study and/or provided data but did not necessarily participate in the analysis or writing of this report.

## Notes

### Competing Interest Statement

The authors have declared no competing interest.

## References

Achenback, T. M., & Rescorla, L. A. (2001). Manual for the ASEBA School-Age Forms & Profiles. In. Burlington, VT: ASEBA.

Albajara Sáenz, A., Villemonteix, T., & Massat, I. (2019). Structural and functional neuroimaging in attention-deficit/hyperactivity disorder. Dev Med Child Neurol, 61(4), 399–405. doi:10.1111/dmcn.14050

American Psychiatric Association., & American Psychiatric Association. DSM-5 Task Force. (2013). Diagnostic and statistical manual of mental disorders: DSM-5 (5th ed.). Arlington, Va.: American Psychiatric Association.

Angulo-Ruiz, B. Y., Muñoz, V., Rodríguez-Martínez, E. I., Cabello-Navarro, C., & Gómez, C. M. (2022). Multiscale entropy of ADHD children during resting state condition. Cognitive Neurodynamics. doi:10.1007/s11571-022-09869-0

Benjamini, Y., & Hochberg, Y. (1995). Controlling the false discovery rate: a practical and powerful approach to multiple testing. Journal of the Royal Statistical Society. Series B (Methodological), 57(1), 12.

Biswal, B., Yetkin, F. Z., Haughton, V. M., & Hyde, J. S. (1995). Functional connectivity in the motor cortex of resting human brain using echo-planar MRI. Magn Reson Med, 34(4), 537–541. doi:10.1002/mrm.1910340409

Buckner, R. L., & Krienen, F. M. (2013). The evolution of distributed association networks in the human brain. Trends Cogn Sci, 17(12), 648–665. doi:10.1016/j.tics.2013.09.017

Casey, B. J., Cannonier, T., Conley, M. I., Cohen, A. O., Barch, D. M., Heitzeg, M. M.,… Workgroup, A. I. A. (2018). The Adolescent Brain Cognitive Development (ABCD) study: Imaging acquisition across 21 sites. Dev Cogn Neurosci, 32, 43–54. doi:10.1016/j.dcn.2018.03.001

Castellanos, F. X., & Aoki, Y. (2016). Intrinsic Functional Connectivity in Attention-Deficit/Hyperactivity Disorder: A Science in Development. Biol Psychiatry Cogn Neurosci Neuroimaging, 1(3), 253–261. doi:10.1016/j.bpsc.2016.03.004

Cortese, S., Aoki, Y. Y., Itahashi, T., Castellanos, F. X., & Eickhoff, S. B. (2021). Systematic Review and Meta-analysis: Resting-State Functional Magnetic Resonance Imaging Studies of Attention-Deficit/Hyperactivity Disorder. J Am Acad Child Adolesc Psychiatry, 60(1), 61–75. doi:10.1016/j.jaac.2020.08.014

Cortese, S., Kelly, C., Chabernaud, C., Proal, E., Di Martino, A., Milham, M. P., & Castellanos, F. X. (2012). Toward systems neuroscience of ADHD: a meta-analysis of 55 fMRI studies. Am J Psychiatry, 169(10), 1038–1055. doi:10.1176/appi.ajp.2012.11101521

Costa, M., Goldberger, A. L., & Peng, C. K. (2002). Multiscale entropy analysis of complex physiologic time series. Phys Rev Lett, 89(6), 068102. doi:10.1103/PhysRevLett.89.068102

Desikan, R. S., Ségonne, F., Fischl, B., Quinn, B. T., Dickerson, B. C., Blacker, D.,… Killiany, R. J. (2006). An automated labeling system for subdividing the human cerebral cortex on MRI scans into gyral based regions of interest. Neuroimage, 31(3), 968–980. doi:10.1016/j.neuroimage.2006.01.021

Fair, D. A., Cohen, A. L., Power, J. D., Dosenbach, N. U., Church, J. A., Miezin, F. M.,… Petersen, S. E. (2009). Functional brain networks develop from a “local to distributed” organization. PLoS Comput Biol, 5(5), e1000381. doi:10.1371/journal.pcbi.1000381

Fernández, A., Gómez, C., Hornero, R., & López-Ibor, J. J. (2013). Complexity and schizophrenia. Prog Neuropsychopharmacol Biol Psychiatry, 45, 267–276. doi:10.1016/j.pnpbp.2012.03.015

Fernández, A., Quintero, J., Hornero, R., Zuluaga, P., Navas, M., Gómez, C.,… Ortiz, T. (2009). Complexity analysis of spontaneous brain activity in attention-deficit/hyperactivity disorder: diagnostic implications. Biol Psychiatry, 65(7), 571–577. doi:10.1016/j.biopsych.2008.10.046

Fox, M. D., Zhang, D., Snyder, A. Z., & Raichle, M. E. (2009). The global signal and observed anticorrelated resting state brain networks. J Neurophysiol, 101(6), 3270–3283. doi:10.1152/jn.90777.2008

Friedman, L. A., & Rapoport, J. L. (2015). Brain development in ADHD. Curr Opin Neurobiol, 30, 106–111. doi:10.1016/j.conb.2014.11.007

Frodl, T., & Skokauskas, N. (2012). Meta-analysis of structural MRI studies in children and adults with attention deficit hyperactivity disorder indicates treatment effects. Acta Psychiatr Scand, 125(2), 114–126. doi:10.1111/j.1600-0447.2011.01786.x

Gao, Y., Shuai, D., Bu, X., Hu, X., Tang, S., Zhang, L.,… Huang, X. (2019). Impairments of large-scale functional networks in attention-deficit/hyperactivity disorder: a meta-analysis of resting-state functional connectivity. Psychol Med, 49(15), 2475–2485. doi:10.1017/S003329171900237X

Gu, C., Liu, Z. X., & Woltering, S. (2022). Electroencephalography complexity in resting and task states in adults with attention-deficit/hyperactivity disorder. Brain Commun, 4(2), fcac054. doi:10.1093/braincomms/fcac054

Guan, S., Wan, D., Zhao, R., Canario, E., Meng, C., & Biswal, B. B. (2023). The complexity of spontaneous brain activity changes in schizophrenia, bipolar disorder, and ADHD was examined using different variations of entropy. Hum Brain Mapp, 44(1), 94–118. doi:10.1002/hbm.26129

Gómez, C., Poza, J., Fernández, A., Bachiller, A., Gómez, J., & Hornero, R. (2013). Entropy analysis of MEG background activity in attention-deficit/hyperactivity disorder. Annu Int Conf IEEE Eng Med Biol Soc, 2013, 5057–5060. doi:10.1109/EMBC.2013.6610685

Hart, H., Radua, J., Nakao, T., Mataix-Cols, D., & Rubia, K. (2013). Meta-analysis of functional magnetic resonance imaging studies of inhibition and attention in attention-deficit/hyperactivity disorder: exploring task-specific, stimulant medication, and age effects. JAMA Psychiatry, 70(2), 185–198. doi:10.1001/jamapsychiatry.2013.277

Hu, Z., Liu, L., Wang, M., Jia, G., Li, H., Si, F.,… Niu, H. (2021). Disrupted signal variability of spontaneous neural activity in children with attention-deficit/hyperactivity disorder. Biomed Opt Express, 12(5), 3037–3049. doi:10.1364/BOE.418921

Kessler, D., Angstadt, M., Welsh, R. C., & Sripada, C. (2014). Modality-spanning deficits in attention-deficit/hyperactivity disorder in functional networks, gray matter, and white matter. J Neurosci, 34(50), 16555–16566. doi:10.1523/JNEUROSCI.3156-14.2014

Kumar, U., Arya, A., & Agarwal, V. (2021). Neural network connectivity in ADHD children: an independent component and functional connectivity analysis of resting state fMRI data. Brain Imaging Behav, 15(1), 157–165. doi:10.1007/s11682-019-00242-0

Lin, H. Y., Kessler, D., Tseng, W. I., & Gau, S. S. (2021). Increased Functional Segregation Related to the Salience Network in Unaffected Siblings of Youths With Attention-Deficit/Hyperactivity Disorder. J Am Acad Child Adolesc Psychiatry, 60(1), 152–165. doi:10.1016/j.jaac.2019.11.012

Liu, N., Liu, Q., Yang, Z., Xu, J., Fu, G., Zhou, Y.,… Qian, Q. (2023). Different functional alteration in attention-deficit/hyperactivity disorder across developmental age groups: A meta-analysis and an independent validation of resting-state functional connectivity studies. CNS Neurosci Ther, 29(1), 60–69. doi:10.1111/cns.14032

Marek, S., & Dosenbach, N. U. F. (2018). The frontoparietal network: function, electrophysiology, and importance of individual precision mapping. Dialogues Clin Neurosci, 20(2), 133–140. doi:10.31887/DCNS.2018.20.2/smarek

McDonough, I. M., & Nashiro, K. (2014). Network complexity as a measure of information processing across resting-state networks: evidence from the Human Connectome Project. Front Hum Neurosci, 8, 409. doi:10.3389/fnhum.2014.00409

Paloyelis, Y., Mehta, M. A., Kuntsi, J., & Asherson, P. (2007). Functional MRI in ADHD: a systematic literature review. Expert Rev Neurother, 7(10), 1337–1356. doi:10.1586/14737175.7.10.1337

Petersen, A. C., Crockett, L., Richards, M., & Boxer, A. (1988). A self-report measure of pubertal status: Reliability, validity, and initial norms. J Youth Adolesc, 17(2), 117–133. doi:10.1007/BF01537962

Pincus, S. M. (1991). Approximate entropy as a measure of system complexity. Proc Natl Acad Sci U S A, 88(6), 2297–2301. doi:10.1073/pnas.88.6.2297

Polanczyk, G., de Lima, M. S., Horta, B. L., Biederman, J., & Rohde, L. A. (2007). The worldwide prevalence of ADHD: a systematic review and metaregression analysis. Am J Psychiatry, 164(6), 942–948. doi:10.1176/ajp.2007.164.6.942

Posner, J., Park, C., & Wang, Z. (2014). Connecting the dots: a review of resting connectivity MRI studies in attention-deficit/hyperactivity disorder. Neuropsychol Rev, 24(1), 3–15. doi:10.1007/s11065-014-9251-z

Posner, J., Rauh, V., Gruber, A., Gat, I., Wang, Z., & Peterson, B. S. (2013). Dissociable attentional and affective circuits in medication-naïve children with attention-deficit/hyperactivity disorder. Psychiatry Res, 213(1), 24–30. doi:10.1016/j.pscychresns.2013.01.004

Power, J. D., Mitra, A., Laumann, T. O., Snyder, A. Z., Schlaggar, B. L., & Petersen, S. E. (2014). Methods to detect, characterize, and remove motion artifact in resting state fMRI. Neuroimage, 84, 320–341. doi:10.1016/j.neuroimage.2013.08.048

Rezaeezadeh, M., Shamekhi, S., & Shamsi, M. (2020). Attention Deficit Hyperactivity Disorder Diagnosis using non-linear univariate and multivariate EEG measurements: a preliminary study. Phys Eng Sci Med, 43(2), 577–592. doi:10.1007/s13246-020-00858-3

Richman, J. S., & Moorman, J. R. (2000). Physiological time-series analysis using approximate entropy and sample entropy. Am J Physiol Heart Circ Physiol, 278(6), H2039–2049. doi:10.1152/ajpheart.2000.278.6.H2039

Rubia, K. (2011). “Cool” inferior frontostriatal dysfunction in attention-deficit/hyperactivity disorder versus “hot” ventromedial orbitofrontal-limbic dysfunction in conduct disorder: a review. Biol Psychiatry, 69(12), e69–87. doi:10.1016/j.biopsych.2010.09.023

Shaw, M., Hodgkins, P., Caci, H., Young, S., Kahle, J., Woods, A. G., & Arnold, L. E. (2012). A systematic review and analysis of long-term outcomes in attention deficit hyperactivity disorder: effects of treatment and non-treatment. BMC Med, 10, 99. doi:10.1186/1741-7015-10-99

Shaw, P., Eckstrand, K., Sharp, W., Blumenthal, J., Lerch, J. P., Greenstein, D.,… Rapoport, J. L. (2007). Attention-deficit/hyperactivity disorder is characterized by a delay in cortical maturation. Proc Natl Acad Sci U S A, 104(49), 19649–19654. doi:10.1073/pnas.0707741104

Shirer, W. R., Ryali, S., Rykhlevskaia, E., Menon, V., & Greicius, M. D. (2012). Decoding subject-driven cognitive states with whole-brain connectivity patterns. Cereb Cortex, 22(1), 158–165. doi:10.1093/cercor/bhr099

Smith, R. X., Yan, L., & Wang, D. J. (2014). Multiple time scale complexity analysis of resting state FMRI. Brain Imaging Behav, 8(2), 284–291. doi:10.1007/s11682-013-9276-6

Sokunbi, M. O., Fung, W., Sawlani, V., Choppin, S., Linden, D. E., & Thome, J. (2013). Resting state fMRI entropy probes complexity of brain activity in adults with ADHD. Psychiatry Res, 214(3), 341–348. doi:10.1016/j.pscychresns.2013.10.001

Sun, J., Wang, B., Niu, Y., Tan, Y., Fan, C., Zhang, N.,… Xiang, J. (2020). Complexity Analysis of EEG, MEG, and fMRI in Mild Cognitive Impairment and Alzheimer’s Disease: A Review. Entropy (Basel), 22(2). doi:10.3390/e22020239

Sutcubasi, B., Metin, B., Kurban, M. K., Metin, Z. E., Beser, B., & Sonuga-Barke, E. (2020). Resting-state network dysconnectivity in ADHD: A system-neuroscience-based meta-analysis. World J Biol Psychiatry, 21(9), 662–672. doi:10.1080/15622975.2020.1775889

Takahashi, T. (2013). Complexity of spontaneous brain activity in mental disorders. Prog Neuropsychopharmacol Biol Psychiatry, 45, 258–266. doi:10.1016/j.pnpbp.2012.05.001

Wang, D. J. J., Jann, K., Fan, C., Qiao, Y., Zang, Y. F., Lu, H., & Yang, Y. (2018). Neurophysiological Basis of Multi-Scale Entropy of Brain Complexity and Its Relationship With Functional Connectivity. Front Neurosci, 12, 352. doi:10.3389/fnins.2018.00352

Wang, Z., Li, Y., Childress, A. R., & Detre, J. A. (2014). Brain entropy mapping using fMRI. PLoS One, 9(3), e89948. doi:10.1371/journal.pone.0089948

Zang, Y. F., He, Y., Zhu, C. Z., Cao, Q. J., Sui, M. Q., Liang, M.,… Wang, Y. F. (2007). Altered baseline brain activity in children with ADHD revealed by resting-state functional MRI. Brain Dev, 29(2), 83–91. doi:10.1016/j.braindev.2006.07.002

