## Supplementary material for "Functional Connectivity and Complexity Analyses of Resting-State fMRI in Pre-Adolescents with ADHD": NA

Follow-up Analyses

A factorial 2 (Medication: medicated ADHD, non-medicated ADHD) by 2 (Sex: Male, Female) ANCOVA was performed on SampEn at each temporal scale in each ROI with PDS z-score and site as covariates. Main effects of Medication were obtained in the left putamen at scale = 9 (RMN; *F* = 4.868, *p* < 0.05, $\eta_{p}^{2}$ = 0.196), left anterior supramarginal gyrus at scale = 9 to 15 (DMN; *F* = 4.868 – 7.759, *p* < 0.05, $\eta_{p}^{2}$ = 0.196 – 0.280), left middle frontal gyrus at scale = 11 to 14 (FPN; *F* = 4.723 – 6.075, *p* < 0.05, $\eta_{p}^{2}$ =0.191 – 0.233), posterior cingulate gyrus at scale = 13 (DMN; *F* = 6.466, *p* < 0.05, $\eta_{p}^{2}$ = 0.244), anterior cingulate gyrus at scale = 13 (FPN & DMN; *F* = 4.896, *p* < 0.05, $\eta_{p}^{2}$ = 0.197), left superior frontal gyrus at scale = 13 (FPN; *F* = 4.818, *p* < 0.05, $\eta_{p}^{2}$ = 0.194), left posterior supramarginal gyrus at scale = 13 (DMN; *F* = 4.388, *p* < 0.05, $\eta_{p}^{2}$ = 0.180), and medial frontal cortex at scale = 13 (DMN: *F* = 4.375, *p* < 0.05, $\eta_{p}^{2}$ = 0.179). However, none of them survived the Benjamini-Hochberg correction (false discovery rate = 0.05).

Table S1. Brain regions displaying significant main effects of Group, obtained from the ANCOVAs for functional connectivity.

| Region^a^ | *F*-value | *t*-value | $\eta_{p}^{2}$ |
| --- | --- | --- | --- |
| **Within FPN** |  |  |  |
| Right middle frontal gyrus-to-left superior frontal gyrus | 4.526 | 1.573 | 0.035 |
| Anterior cingulate gyrus-to-right superior frontal gyrus | 6.852 | 1.801 | 0.052 |
| Anterior cingulate gyrus-to-left superior frontal gyrus | 5.564 | 1.840 | 0.042 |
| Anterior cingulate gyrus-to-left middle frontal gyrus | 7.052 | 3.007 | 0.053 |
| **Within DMN** |  |  |  |
| Medial frontal cortex-to-right posterior supramarginal gyrus | 4.082 | 1.998 | 0.031 |
| **Within RMN** |  |  |  |
| Right frontal orbital cortex-to-right caudate | 5.643 | 1.488 | 0.043 |
| Right frontal orbital cortex-to-left caudate | 8.542 | 2.627 | 0.063 |
| Right frontal orbital cortex-to-right putamen | 4.544 | 2.153 | 0.035 |
| **Between networks** |  |  |  |
| Right superior frontal gyrus-to-medial frontal cortex (FPN-to-DMN) | 4.607 | 2.198 | 0.035 |
| Right middle frontal gyrus-to-posterior cingulate gyrus (FPN-to-DMN) | 4.540 | 0.884 | 0.035 |
| Anterior cingulate gyrus-to-right frontal orbital cortex (FPN/DMN-to-RMN) | 4.265 | 2.032 | 0.033 |
| Right posterior supramarginal gyrus-to-right frontal orbital cortex (DMN-to-RMN) | 4.072 | 0.731 | 0.031 |
| Medial frontal cortex-to-left caudate (DMN-to-RMN) | 7.800 | 3.929 | 0.058 |

Key to Table S1: ^a^ According to the Harvard-Oxford Atlas (Desikan et al., 2006). All results presented at *p* < 0.05, uncorrected. $\eta_{p}^{2}$ = partial eta squared.

Table S2. Brain regions displaying significant group differences, obtained from the group comparison for partial correlation between MSE and functional connectivity.

| Region^a^ | *Z* value |
| --- | --- |
| **Within DMN** |  |
| Right anterior supramarginal gyrus, right anterior supramarginal gyrus-to-medial frontal cortex (DMN, DMN-to-DMN) | -2.224 |
| Left anterior supramarginal gyrus, left anterior supramarginal gyrus-to-medial frontal cortex (DMN, DMN-to-DMN) | -2.075 |
| Medial frontal cortex, medial frontal cortex-to-right anterior supramarginal gyrus (DMN, DMN-to-DMN) | -2.428 |
| Medial frontal cortex, medial frontal cortex-to-left anterior supramarginal gyrus (DMN, DMN-to-DMN) | -2.181 |
| **Between networks** |  |
| Left middle frontal gyrus, left middle frontal gyrus-to-right anterior supramarginal gyrus (FPN, FPN-to-DMN) | 2.239 |
| Left posterior supramarginal gyrus, left posterior supramarginal gyrus-to-right putamen (DMN, DMN-to-RMN) | -2.010 |
| Frontal orbital cortex, frontal orbital cortex-to-left superior frontal gyrus (RMN, RMN-to-PFN) | -2.235 |
| Right caudate, right caudate-to-posterior cingulate gyrus (RMN, RMN-to-DMN) | -2.106 |
| Right putamen, right putamen-to-left posterior supramarginal gyrus (RMN, RMN-to-DMN) | -2.242 |
| Left putamen, left putamen-to- left posterior supramarginal gyrus (RMN, RMN-to-DMN) | -2.217 |

Key to Table S2: ^a^ According to the Harvard-Oxford Atlas (Desikan et al., 2006). All results presented at *p* < 0.05, uncorrected.

Table S3. A list of ADHD prescriptions.

Prescription

Adderall

Azstarys

Serdexmethylphenidate

Dexmethylphendiate

Concerta

Methylphenidate

Focalin

Intuniv

Guanfacine

Gelbree

Viloxazine

Ritalin

Strattera

Atomoxetine

Vyvanse

Adzenys

Evekeo

Daytrana

Quillivant

Amphetamine

Clonidine

Metadate

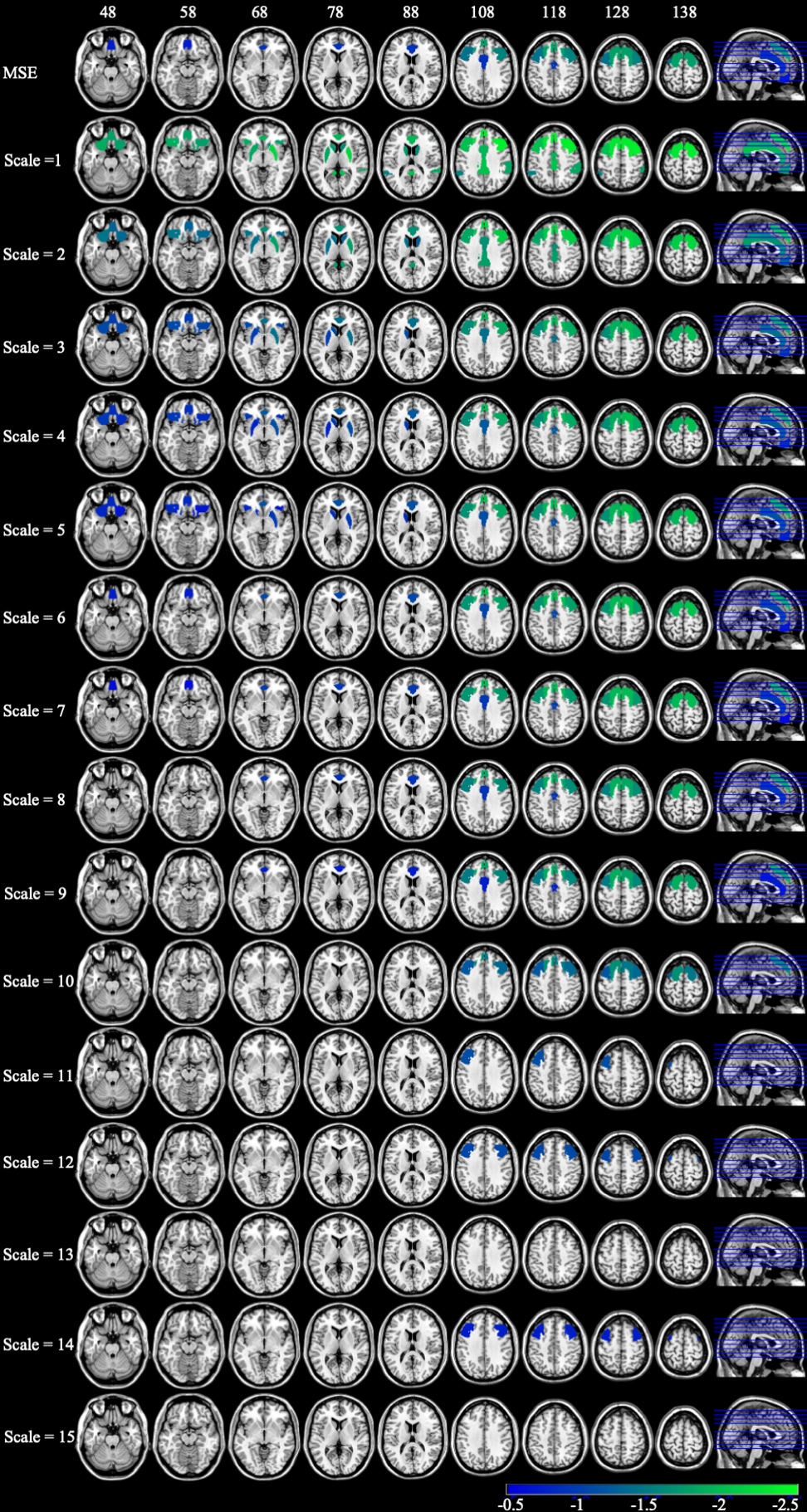

Figure S1. *t* maps of MSE and SampEn at each temporal scale for ADHD vs. control. Only significant main effects of Group are displayed (*p* < 0.05, uncorrected).

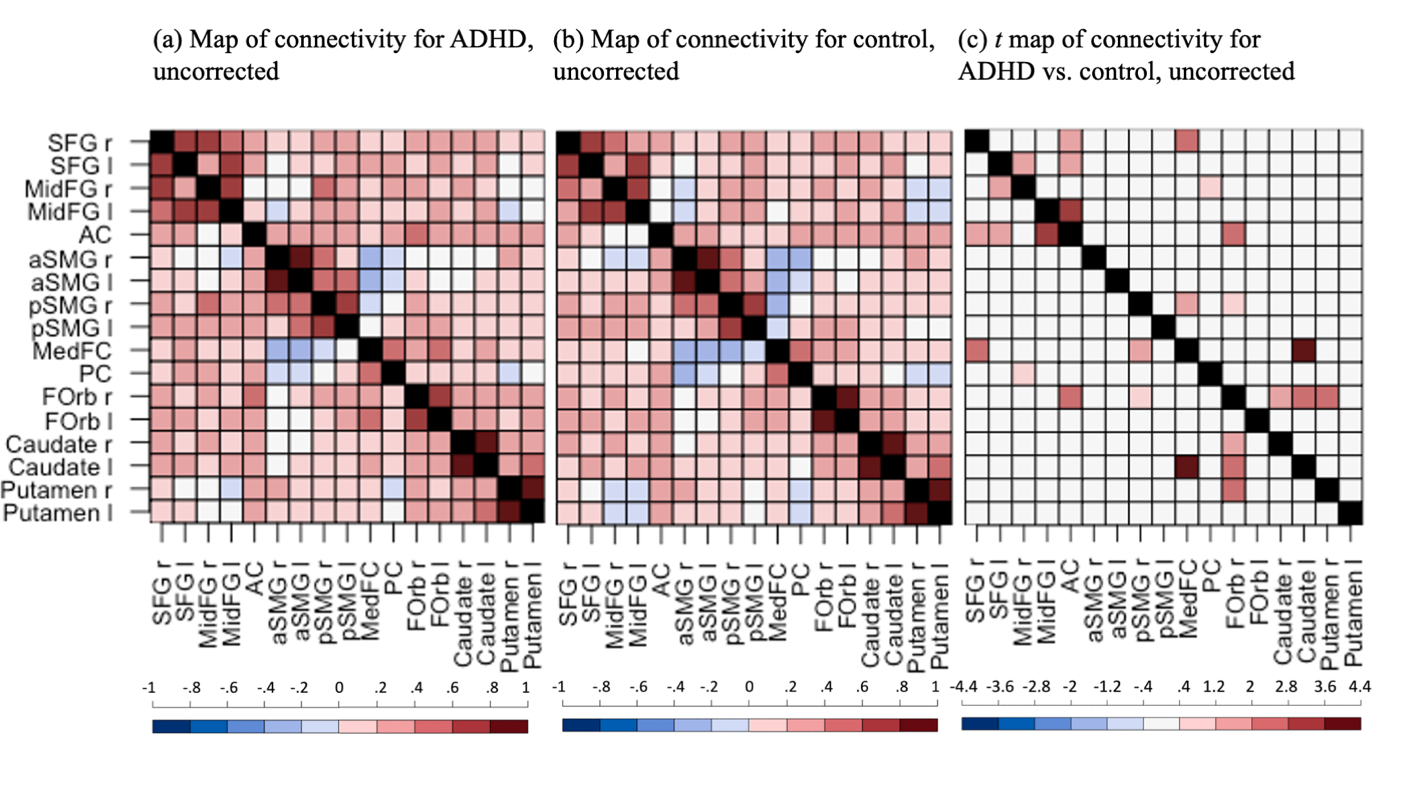
 Figure S2. Connectivity maps (Fisher’s z) for (a) the ADHD group and (b) the control group, and (c) the *t*-value map for the ADHD group vs. the control group. Only significant edges are displayed in blue or red colors, (*p* < 0.05, uncorrected). For (a) and (b), blue/red indicates *p* < 0.05 for the *t*-test against 0. For (c), blue/red indicates *p* < 0.05 for the main effect of Group. The black indicates data was not available. SFG: superior frontal gyrus; MidFG: middle frontal gyrus; AC: anterior cingulate gyrus; aSMG: anterior supramarginal gyrus; pSMG: posterior supramarginal gyrus; MedFC: frontal medial cortex; PC: posterior cingulate gyrus; FOrb: frontal orbital cortex; r: right; l: left.

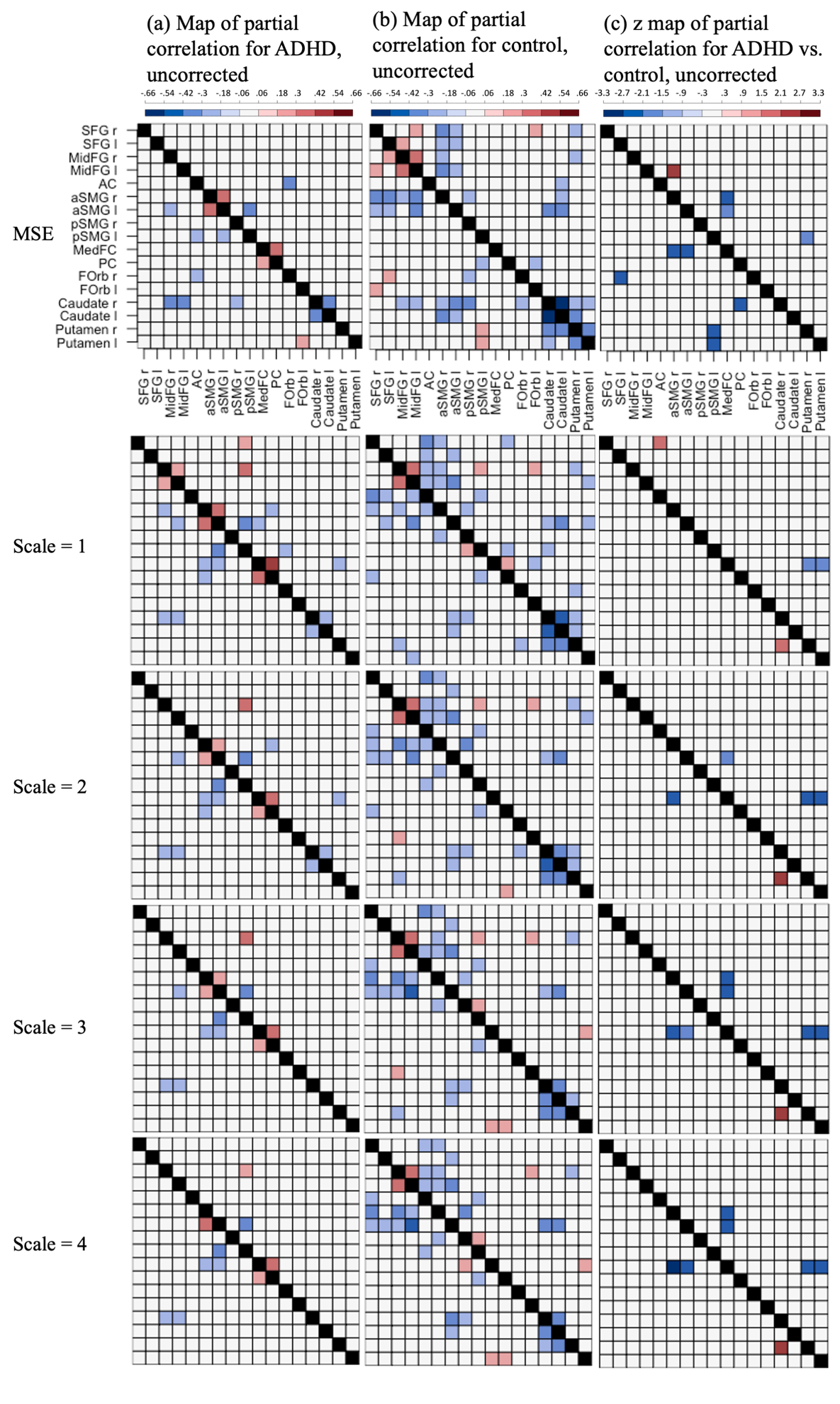

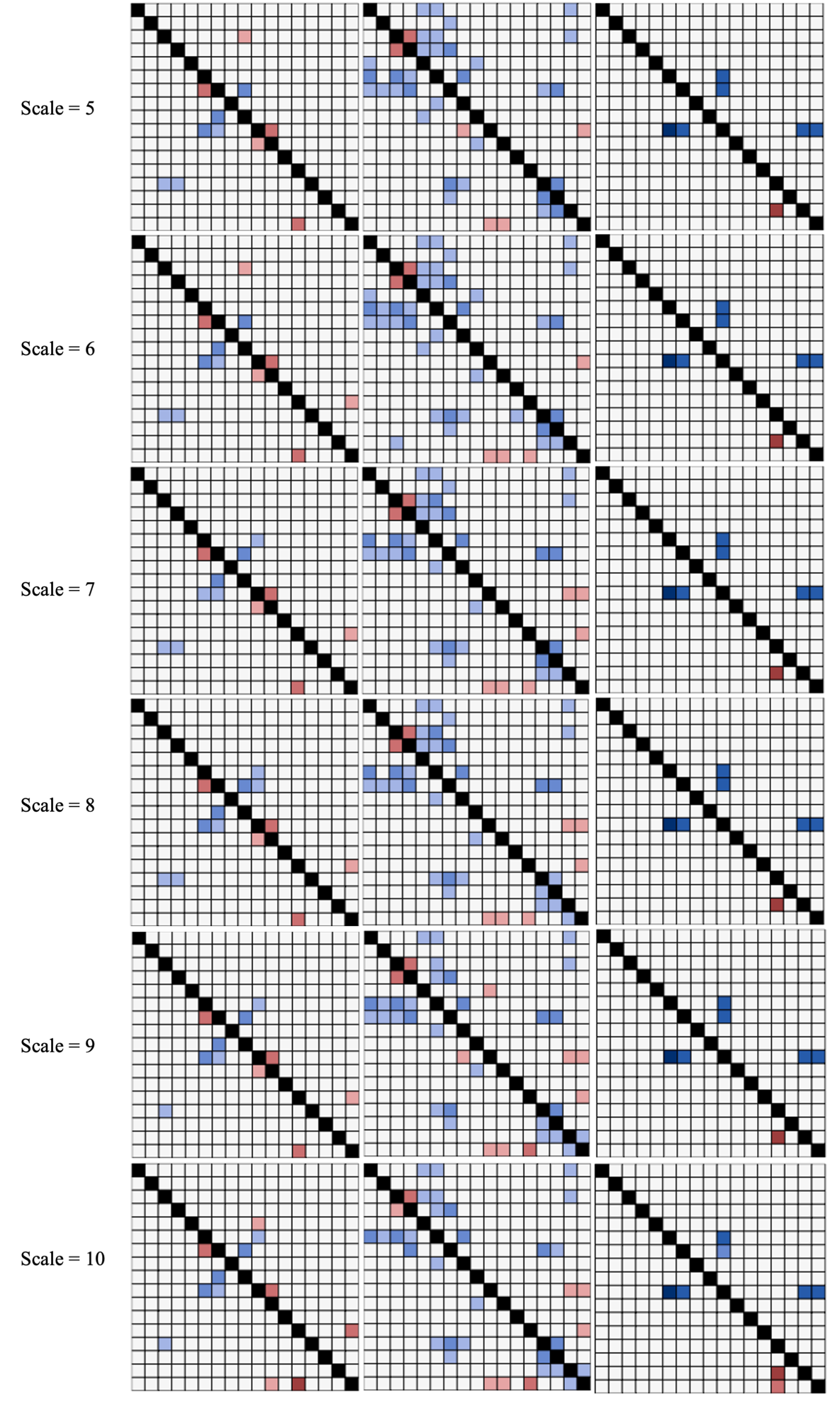

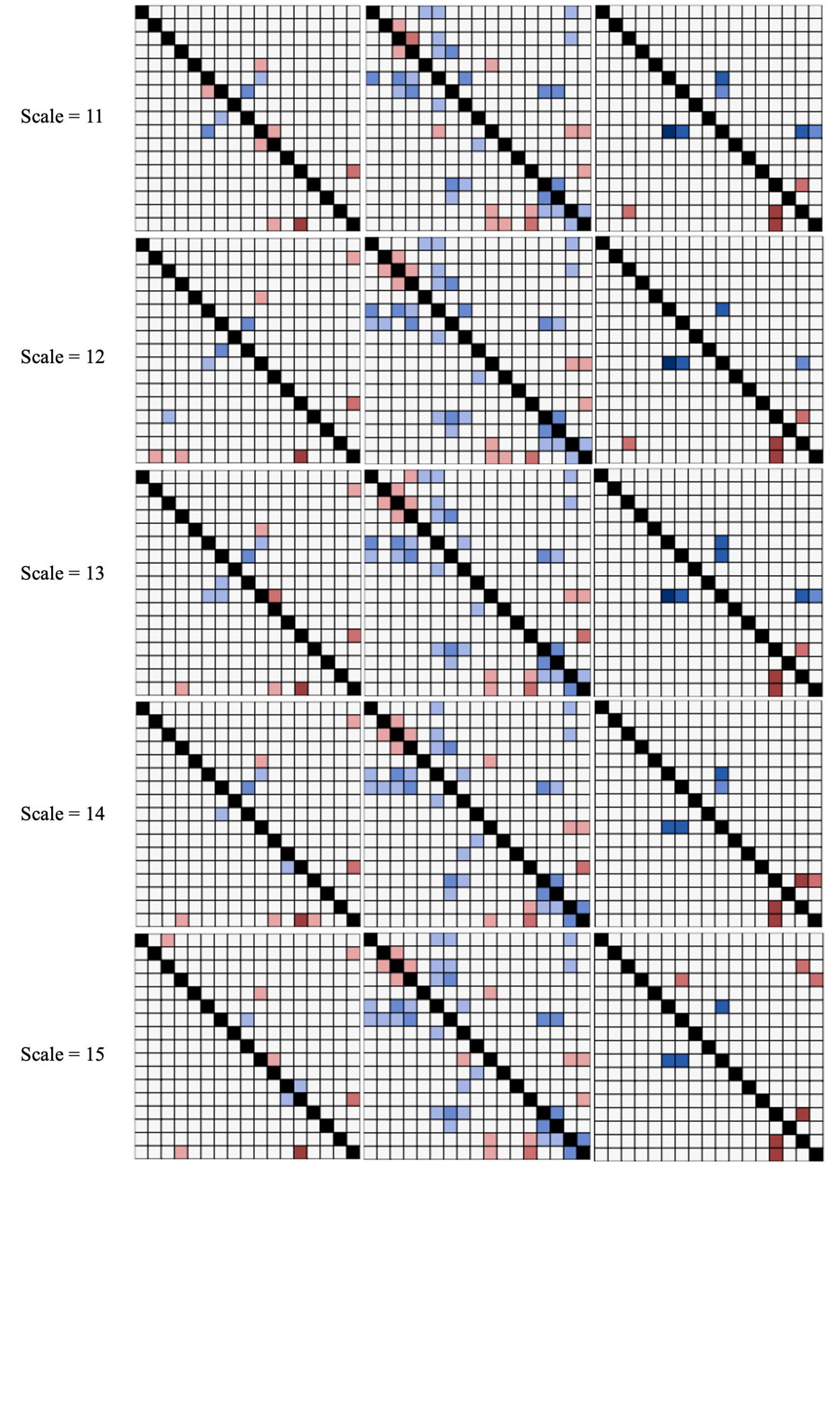

Figure S3. Partial correlation (Fisher’s Z) between MSE or SampEn at different temporal scales and connectivity for (a) the ADHD group and (b) the control group, and (c) z value map of partial correlations for ADHD vs. control. Only significant linkages are displayed in blue or red colors (*p* < 0.05, uncorrected). For (a) and (b), blue/red indicates *p* < 0.05 for the *t*-test against 0. For (c), blue/red indicates *p* < 0.05 for the statistical comparison of partial correlations in the ADHD and control groups. The black indicates data was not available. SFG: superior frontal gyrus; MidFG: middle frontal gyrus; AC: anterior cingulate gyrus; aSMG: anterior supramarginal gyrus; pSMG: posterior supramarginal gyrus; MedFC: frontal medial cortex; PC: posterior cingulate gyrus; FOrb: frontal orbital cortex; r: right; l: left.

References

Desikan, R. S., Ségonne, F., Fischl, B., Quinn, B. T., Dickerson, B. C., Blacker, D., . . . Killiany, R. J. (2006). An automated labeling system for subdividing the human cerebral cortex on MRI scans into gyral based regions of interest. *Neuroimage, 31*(3), 968-980. doi:10.1016/j.neuroimage.2006.01.021
